# Universal epidemic curve for COVID-19 and its usage for forecasting

**DOI:** 10.1101/2020.11.07.20220392

**Authors:** Aryan Sharma, Srujan Sapkal, Mahendra K Verma

**Affiliations:** Department of Physics, Indian Institute of Technology Kanpur, Kanpur 208016, India; Department of Materials Engineering, Defence Institute of, Advanced Technology, Pune 411025, India

**Keywords:** COVID-19, Universal Curve, Epidemic forecast, India’s COVID-19 evolution

## Abstract

We construct a universal epidemic curve for COVID-19 using the epidemic curves of eight nations that have reached saturation for the first phase, and then fit an eight-degree polynomial that passes through the universal curve. We take India’s epidemic curve up to September 22, 2020 and overlap it with the universal curve by minimizing square-root error. The constructed curve is used to forecast epidemic evolution up to January 1, 2021. The predictions of our model and those of supermodel for India are reasonably close to each other considering the uncertainties in data fitting.

## 1 Introduction

The COVID-19 pandemic is one of the most devastating natural disaster in the last 100 years and it is still raging around the world [52]. As of October 25, 2020, the total infection count is around 43 million and the total death cases crossed 1.1 million [51]. The whole world is engaged in mitigation efforts of COVID-19. An important activity in this direction is the prediction of epidemic evolution. Towards this aim, in this paper, we construct a universal epidemic curve for COVID-19 and use this curve to forecast the epidemic evolution in India.

The SIR model. constructed by Kermack and McKendrick [21], is one of the first models for epidemic evolution. In this model, the variables *S* and *I* describe respectively the numbers of susceptible and infected individuals, while the variable *R* represents the removed individuals who have either recovered or died. SEIR model, which is a generalization of SIR mode, includes *exposed* individuals, *E*, who are infected but not yet infectious [6, 14]. Researchers have constructed derivatives of the above models to include lockdowns and travel restrictions, asymptomatic infections, etc. For example, Peng et al. [34] constructed a seven-variable model that includes quarantined and death variables and predicted that the daily count of exposed and infectious individuals in China will be negligible by March 30, 2020. Chinazzi et al. [12] and Hellewell et al. [19] studied the effects of travel restrictions and isolation on epidemic evolution. Mandal et al. [29] constructed a India-specific model that includes intercity connectivity. Shayak et al. [41] modelled epidemic evolution using delayed-differential equations. In addition, Rahmandad et al. [35] has also used a model to predict Indian epidemic growth. Schu‥ttler et al. [39] showed that *I*(*t*) or total death count could be modelled using the error function.

Asymptomatic carriers play a major role in the spread of COVID-19 epidemic, hence there have been many attempts to model this effect. In particular, Ansumali et al. [2] and Robinson et al. [38] have created SAIR model that takes into this important factor. Recently, Vidyasagar at al. [49] and Agrawal et al. [1] have adopted SAIR model to construct an epidemic evolution for India; this model, termed as *supermodel*, has many predictions. For example, it predicts 10.6 million cases by the end of this year.

Data-driven models are also used for epidemic forecast. Recent analysis of COVID-19 data reveals that the epidemic curve begins with an exponential growth, after which we it follows a sequence of power laws [53, 22, 28, 7, 11, 10, 48, 31, 42, 3]. The epidemic curve flattens after square-root growth. Motivated by this observation, in this paper we construct a universal epidemic curve for COVID-19 by appropriate normalization. We use the epidemic data of the first phases of epidemic growth.

The above universal behaviour [28, 32] can be utilized for the predictions of epidemic in various countries. In this paper we overlap India’s epidemic curve on the universal curve by appropriate normalization. We observe that the model predictions describe the past data quite well. In particular, the model forecast for last five weeks are in good agreement with the observed data within 12.3%.

In the next section we construct the universal epidemic curve using the epidemic data of several countries.

### 2 Construction of the universal epidemic curve

To construct the universal curve for the COVID-19 epidemic, we take the epidemic evolution curves of eight countries: France, Spain, Italy, Switzerland, Turkey, Netherlands, Belgium and Germany. We chose these countries because they have reached saturation for the first phase. We obtained the data from ‘EU Open Data Portal’ [13] and WorldOMeter [51] websites. The starting dates of the data collection for these countries are given in Table 1. We take the same end date, 30 June 2020, for all the nations.

**Table 1.** *I*_max_ and *t*_max_ for the eight nations used for the construction of universal curve, the starting date taken for universal curve. The end date for all of them are taken to be June 30, 2020.

| Countries<br>(Start Date) | $I_{\max}$ | $t_{\max}$ |
| --- | --- | --- |
| France<br>(February 24) | 164,801 | 128 |
| Spain<br>(February 26) | 296,351 | 126 |
| Italy<br>(February 21) | 240,578 | 131 |
| Switzerland<br>(February 26) | 31,714 | 126 |
| Turkey<br>(March 12) | 199,906 | 111 |
| Netherlands<br>(February 27) | 50,273 | 125 |
| Belgium<br>(February 29) | 61,427 | 123 |
| Germany<br>(February 23) | 195,832 | 129 |

We consider the curves for cumulative infection counts (*I*(*t*)) versus number of days (*t*). To construct the universal curve, we normalize the curves for the selected countries by dividing *I*(*t*) and *t* with *I*_max_ and *t*_max_ respectively. *I*_max_ and *t*_max_ for each country are defined as the value of *I*(*t*) and *t* as on 30th June 2020 (see Table 1). The normalized *I*(*t*) curves indeed exhibit a universal behaviour, as shown in Fig. 1. The dashed lines represent individual countries, whereas the solid black curve represents the average of all the eight countries.

**Fig 1.**
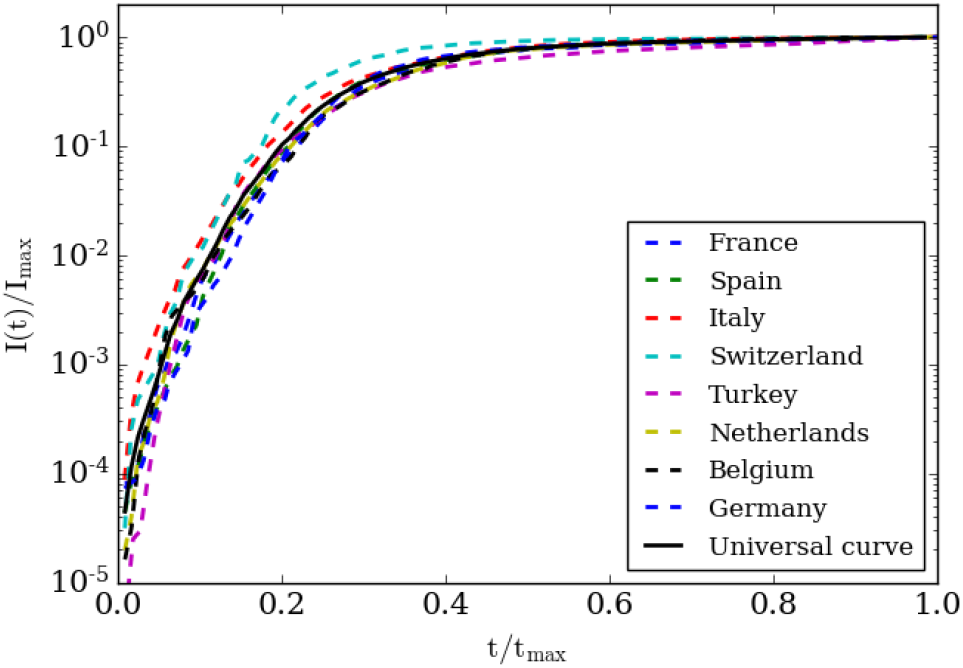
The normalised *I*(*t*) vs. *t* plots of eight countries considered for the construction of the universal curves. The solid black curve represents the average of all the plots.

Note that the universal curve starts with an exponential part and then it contains various power-law regimes before reaching a saturation. Refer to Fig. 2 and references [10, 48, 7, 31, 42, 3] for more details on various power-law regimes of the epidemic curves. Rather than fitting with various power laws at different stages of the epidemic, we fit a large-degree polynomial that passes through the universal curve after the exponential regime. This polynomial is listed in Table 2.

**Table 2.** Polynomial that fit with the universal curve of Fig. 1 and 2.

| Figure Details | Best-fit functions with errors |
| --- | --- |
| A polynomial that fits with the universal curve of Fig. 1 | 1) $-240.5t^8 + 1130t^7 - 2194t^6 + 2253t^5 - 1288t^4 + 389t^3 - 50.26t^2 + 2.539t - 0.03237$ ($std : \pm 0.089$) |
| Best-fit curves for various regimes shown in Fig. 2 | 1) $8.31e^{0.44t} (\pm 3.81\%)$<br>2) $44.54e^{0.24t} (\pm 5.35\%)$<br>3) $3228.69t - 63346.48 (\pm 2.26\%)$<br>4) $14745.83\sqrt{t} + 3354.79 (\pm 1.55\%)$ |

**Fig 2.**
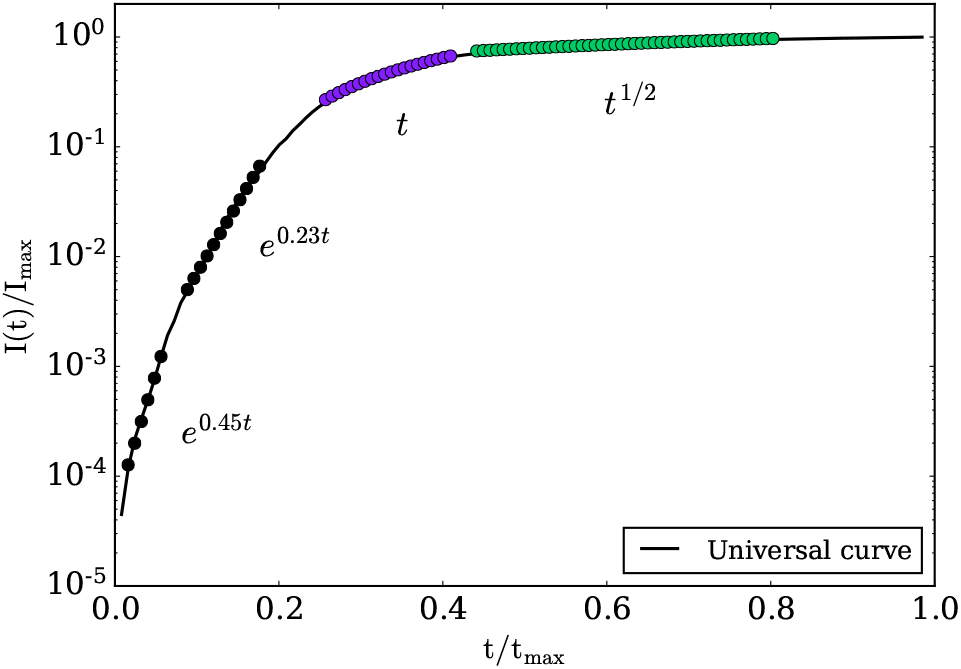
The black curve is the universal curve of Fig. 1. We illustrate various power-law regimes in the universal curve.

**Fig 3.**
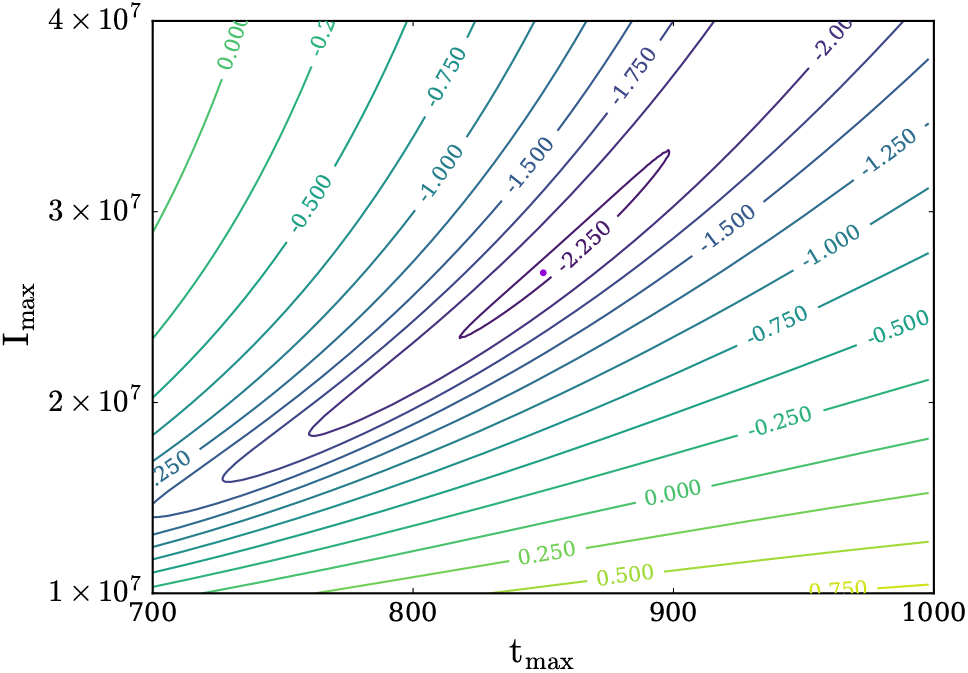
For India’s epidemic curve, contour plot of log(Error) as a function of I_max_ and t_max_, where Error is as defined in Eq. 1. The small dot inside the innermost closed curve represents the minima for which I_max_ = 26.85 million cases and t_max_ = 850 days.

## 3 Modelling Indian epidemic curve using the universal curve

After the construction of an unversal epidemic curve for COVID-19, we attempt to overlap the Indian epidemic curve on the universal curve by employing appropriate normalization. The real-time data of India’s Infection count were accessed from ‘EU Open Data Portal’ from 4th March to 5th October. Note that India’s *I*(*t*) curve is yet to reach saturation, hence we cannot determine *t*_max_ and *I*_max_ from India’s epidemic curve at present. For optimization purpose of our algorithm, we used India’s data from 4th March to 22nd September, 2020. We determine these quantities approximately using the procedure outlined in Algorithm 1.

We estimate *t*_max_ and *I*_max_ by minimizing the following function:

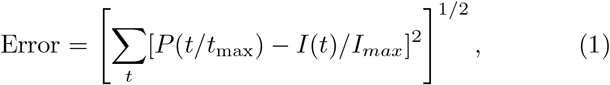

where *P* (*t/t*_max_) is the polynomial fit for the universal function, shown in Table 2. We choose *t*_max_ and *I*_max_ for which the error is minimum (see Algorithm 1).

Fortunately, the process converges towards the unique minimum. The numerical procedure yields *I*_max_ = 26.85 million and *t*_max_ = 850 days for which the value of error in equation 1 is 0.1023. These values yield a maximum overlap for India’s normalised curve on the Universal curve (Fig. 4). We expect to get a better fit with more data (after later date).

### Algorithm 1: Optimizing I_max_ and t_max_.

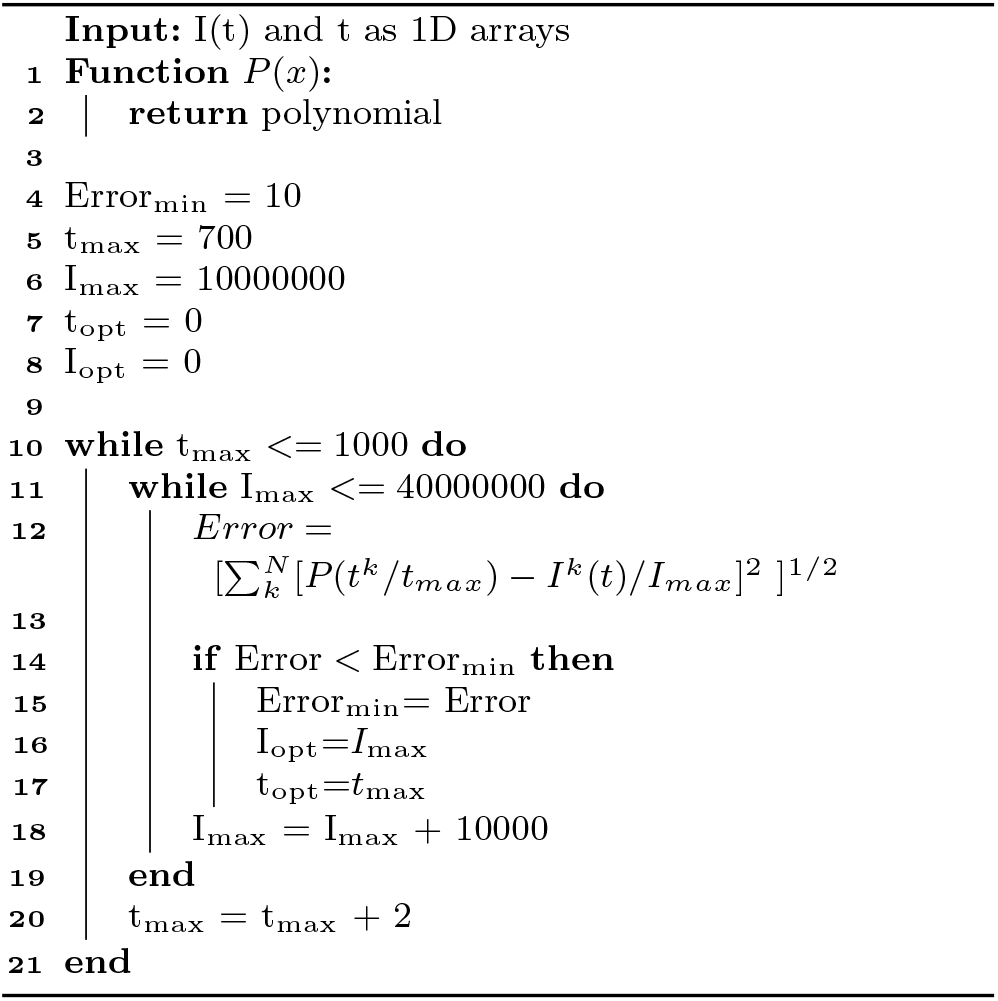

**Fig 4.**
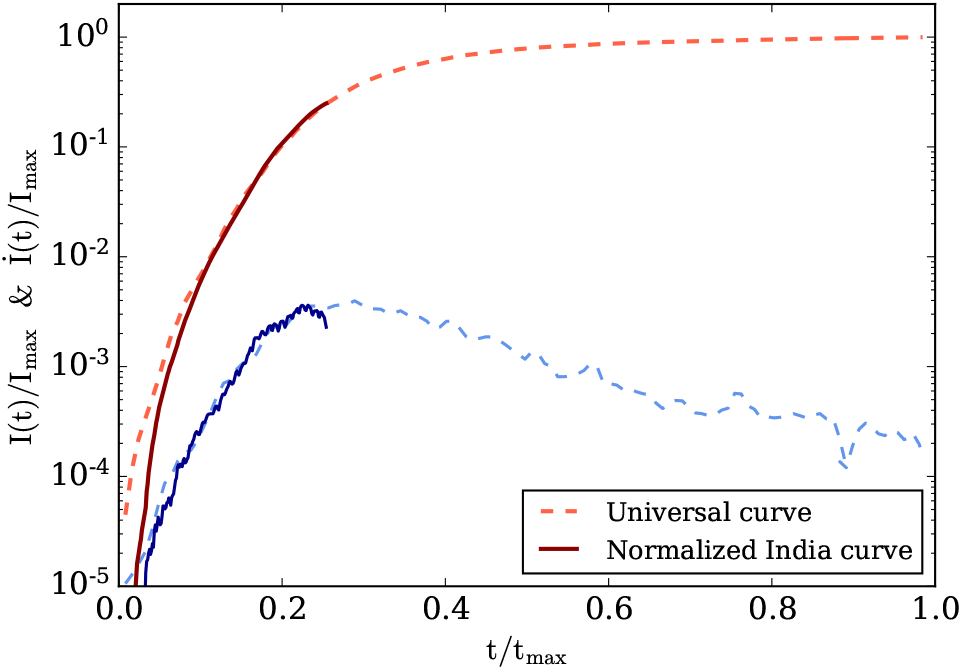
Normalized cumulative (red) and daily infection (blue) counts for India. The solid curves represent the actual data, while the dashed curves are the universal curves using which we can forecast India’s curves.

After this exercise, we construct India’s epidemic curve *I*(*t*) using the universal curve. By multiplying *t/t*_max_ by *t*_max_ and *I*(*t*)*/I*_max_ by *I*_max_, we obtain the predictions for *I*(*t*) for any date, as shown in Fig. 5. We construct a curve for *I* (*t*), which is the daily cases, by taking a numerical derivative of *I*(*t*) curve.

**Fig 5.**
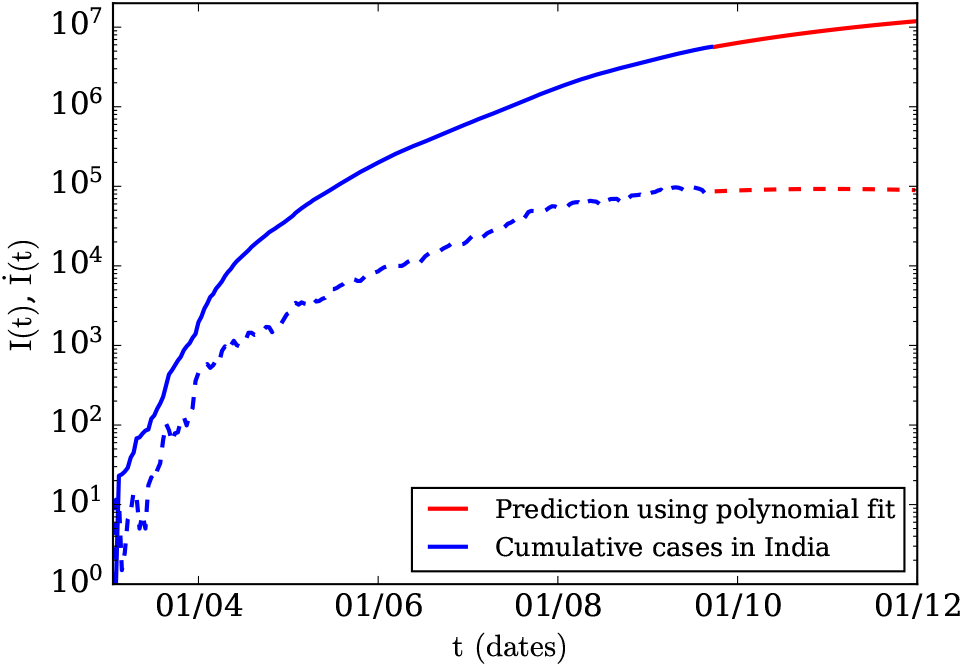
Projections of COVID-19 epidemic curves for India using polynomial fit curve of Table 2.

## 4 Comparing the predictions with the epidemic data

Once India’s epidemic curve *I*(*t*) has been constructed, we can forecast infection count at any date, as shown in Fig. 4 and Fig. 5. Note that the India’s *I*(*t*) curve overlaps with the universal curve quite well. Regarding the forecast, the universal curves indicate that the linear regime starts at around *t/t*_max_ =0.25. For India, this time translates to the last week of September. Note that the daily cases are approximately constant in the linear regime, but they start to decrease after the linear regime.

In Table 3, we list weekly new cases, along with the model predictions, for India. We also illustrate these numbers in Fig. 6. The model predictions are close to the actual data, with the maximum error if 12.3%. A closer look however reveals that the model’s peak for the daily cases is delayed compared to the actual cases. This is somewhat expected due to uncertainties in determination of the universal curve. On the whole, our model predictions appear to be reasonably robust.

**Table 3.** Predictions of new COVID-19 cases on weekly basis using the universal curve or the best-fit polynomial.

| Week | Actual weekly new cases (in millions) | Predicted cases with percentage errors (in millions) |
| --- | --- | --- |
| India: Week-I (Sept 1 - Sept 7) | 0.589 | 0.549 (6.79%) |
| India: Week-II (Sept 8 - Sept 14) | 0.65 | 0.573 (11.85%) |
| India: Week-III (Sept 15 - Sept 21) | 0.632 | 0.593 (6.17%) |
| India: Week-IV (Sept 22 - Sept 28) | 0.583 | 0.612 (4.97%) |
| India: Week-V (Sept 29 - Oct 5) | 0.549 | 0.626 (12.3%) |
| India: Week-VI (Oct 6 - Oct 12) | NA | 0.636 |
| India: Week-VII (Oct 13 - Oct 19) | NA | 0.643 |
| India: Week-VIII (Oct 20 - Oct 26) | NA | 0.647 |
| India: Week-IX (Oct 27 - Nov 2) | NA | 0.649 |
| India: Week-X (Nov 3 - Nov 9) | NA | 0.648 |
| India: Week-XI (Nov 10 - Nov 16) | NA | 0.644 |
| India: Week-XII (Nov 17 - Nov 23) | NA | 0.639 |

**Fig 6.**
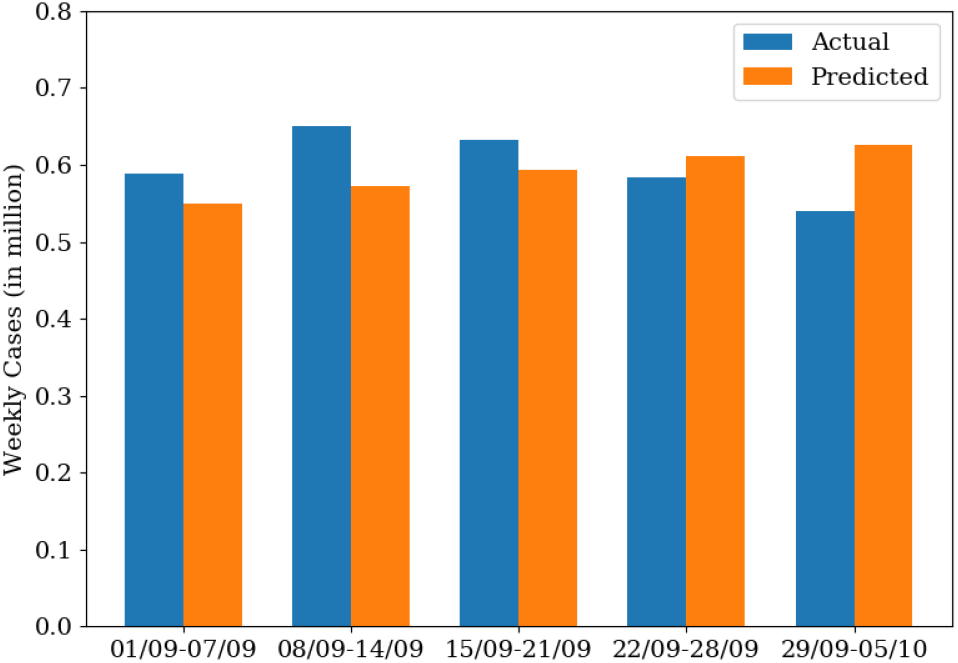
Bar chart of the weekly new cases of India’s epidemic (also see Table 3).

## 5 Comparison with other leading epidemic models for India

As described in the introduction, there are interesting low-dimension models of epidemic evolution. These models are refinement of SEIR model. In this section, we compare our model predictions with some of the leading epidemic models for India. In one such model, Rahmandad et al. [35] forecasted that in early 2021, the daily infections count in India will reach 0.287 million (2.87 lacs). Also, refer to Song et al. [43]. Our model predicts that the daily count will much lower than the above numbers, unless India faces a devastating second wave.

India’s supermodel [49, 1], which is based on SAIR model [2, 38], has gained major prominence recently. This model predicts that India may have reached herd immunity with around 38 crores of the population either infected or having antibodies. One of the predictions of the supermodel is that the infection counts at the end 2021 would be 10.6 million. For January 1, 2021, our prediction for the total infection count is approximately 14.57 million, and that for daily cont is 81 thousands. These are over-predictions compared to supermodel, but the predictions are reasonably close. We believe that our present model could be improved significantly by employing machine learning algorithms.

## 6 Discussions and Conclusions

We construct a universal epidemic curve for COVID-19 using the first-phase epidemic curves for the eight countries: France, Spain, Italy, Switzerland, Turkey, Netherlands, Belgium and Germany. The curves for the individual countries collapse to a single curve within a standard deviation of 0.089 indicating a generic behaviour of the epidemic. We also construct a eight-degree polynomial that fits with the universal curve.

Universality of the epidemic curve is an important landmark considering that many major physical phenomena exhibit universality (e.g., law of gravitation, phase transition, etc.). Note however that the universality in epidemic is somewhat surprising considering major differences in demography, government actions, lockdown conditions, etc.

The discovery of the universal epidemic curve gives us an interesting handle for forecasting the epidemic evolution. An advantage of this approach over others is that it is purely data-driven. Hence, we do not need to model the differential equation. A disadvantage of this method is that we do not have any control parameter. For example, SAIR model can be tuned by changing the coefficient of some terms of the differential equations, but we cannot do so in our model because we do not have any control over the data.

We compared India’s reported epidemic curve with the universal curve with appropriate scaling. We observed that India’s present epidemic curve fits with the part of the universal curve. This discovery enables us to forecast the epidemic evolution. We observe that our forecasts for 5 weeks match with the observed data within 12.3%, which is quite encouraging considering so many uncertainties. Note however that our predictions tend to be systematically larger than the actual data, which could be due to errors in the construction of the universal curve.

Our model predicts that the daily cases for India’s COVID-19 epidemic are falling, which is consistent with the observations. This result indicates the cumulative *I*(*t*) has reached a linear regime for India’s epidemic. The predictions of our models and those of the supermodel are reasonably close to each other.

The universal curve could be further refined using more advanced algorithm, such as machine learning and deep neural networks. Also, we believe that this model is robust for modelling the second and third waves of COVID-19 as well. In addition, it will be interesting to work out the universal curves for the daily cases, as well as for the active cases. We are in the process of such extensions.

## Data Availability

We have used public data (e.g., from worldometer.org). Paper-spectific plot data will be available on request.

## Acknowledgements

We thank Soumyadeep Chatterjee, Shashwat Bhattacharya, and Asad Ali for useful discussions. This project is supported by a SERB MATRICS project SERB/F/847/2020-2021.

## Notes

### Competing Interest Statement

The authors have declared no competing interest.

### Clinical Trial

N/A

### Author Declarations

We have worked on epidemic evolution of COVID-19 using public data. We have not performed clinical trials on humans. Hence approval from IRB/oversight body is not applicable. We declare that we have followed all scientiif ethics in our research work.

